# A feasibility study examining the impact of an Instagram intervention to improve awareness of antimicrobial resistance among undergraduate students

**DOI:** 10.1101/2025.07.13.25331473

**Authors:** Sana Parveen, Patricia McHugh, Aswathi Surendran, James Matthews, Akke Vellinga

## Abstract

**Background:** Social media offers opportunities to deliver and disseminate health interventions. Social media campaigns have been found to be effective in raising public awareness of health issues during the COVID-19 pandemic but little social media is used to raise awareness on antimicrobial resistance (AMR).

**Objective:** To study the potential of Instagram as a tool for promoting awareness and understanding of AMR among undergraduate students.

**Methods and Analysis:** A 3-month Instagram-based educational intervention with messages about AMR was implemented from January to April 2024 guided by the Transtheoretical Model (TTM). The planning phase involved two co-design sessions and two key-informant interviews which informed the AMR messages. To evaluate the impact of the intervention, pre- and post-intervention surveys were conducted to measure knowledge levels of students. A focus group was also conducted six months post-intervention to assess impacts of the social media intervention. Engagement was calculated by the follower count and reach of the post.

**Results:** 402 students answered the pre-intervention survey and 78 answered the post-intervention survey. 128 students signed up for the intervention by following the Instagram account. A total of 36 messages were posted during the intervention with an overall engagement of 2% by reach. The average likes per post was 14. The co-design sessions and key-informant interviews highlighted visual appeal and content clarity as most important with a preference for simple and concise health information, formal language and user interaction through quizzes. Our findings demonstrate that awareness-raising alone is insufficient and the initial development of understanding and motivation through the early-stage processes of change must be supported by mechanisms which enhance commitment, habit formation and environmental reinforcement.

**Conclusion:** A multi-channel planned intervention targeting the different stages of change is needed to ensure long-term knowledge retention.

## Introduction

Antimicrobial resistance (AMR) is a critical global public health challenge, threatening the effectiveness of antibiotics used to treat infections [1]. According to the World Health Organization (WHO), AMR is identified as one of the ten most severe global health threats, warning that improper antibiotic use and overuse leads to the emergence of resistant bacterial strains [2]. The existing knowledge gaps about prudent antibiotic usage require innovative and targeted educational interventions to reach at-risk populations effectively [3].

Social media enable fast public health message dissemination which can lead to increased public reach and engagement [4, 5]. Research demonstrates that young adults are increasingly depending on Instagram, TikTok and Twitter to obtain health information [6, 7]. Social media facilitates sharing through visual and interactive features which makes them a promising tool for public health education [8, 9]. Given that young adults are among the most active social media users [10], leveraging these platforms to promote responsible antibiotic use and AMR awareness could yield important public health benefits. However, there is little evidence on the effectiveness of social media-based interventions for AMR education [11].

A systematic review suggested educating and training university students could be a good strategy as this cohort of young adults are at a critical stage of forming independent health behaviours and, as future parents, they have the potential to influence public attitudes and practices regarding antibiotic use [11, 12]. However, knowledge about AMR among university students shows wide variations reflected in misconceptions about antibiotics’ effectiveness and proper usage [13, 14]. University students’ high engagement with digital content makes a tailored social media-based interventions a potentially effective method to improve their understanding of AMR and provide evidence-based information.

There is some evidence that theory-based health interventions are more effective than those that are not grounded in theory. One such theory is the Transtheoretical Model (TTM), which offers a comprehensive framework for understanding how individuals progress through different stages of behaviour change [15]. The model integrates both cognitive and behavioural processes, recognising that change does not occur in a single step but through a cyclical progression of stages. In this study, we are not measuring a change in behaviour but rather focusing on the cognitive processes including knowledge, attitudes and intention that are proposed to influence behaviour. Therefore, the intervention employs strategies specifically designed to target early stage cognitive processes of change. Additionally, a co-design approach was used in the development of the intervention involving students in content development process. This ensured the relevance, relatability and acceptability of the intervention to the target audience.

This study aimed to develop, implement and evaluate a three month Instagram based intervention. The target audience was first and second year undergraduate students. The goal was to improve their knowledge, attitudes and intentions related to antimicrobial resistance (AMR) through engaging, evidence-based social media content. In addition to evaluating the effectiveness of the intervention, the study also explored key aspects of feasibility, including the practicality, acceptability, recruitment and retention of participants factors highlighted in the UK MRC framework for complex interventions [16]. The evaluation strategy uses multiple approaches to assess both short- and longer-term effects of the intervention.. This study aimed to explore 3 research questions:

1. Is Instagram an effective tool to promote awareness and understanding of AMR among undergraduate students.
2. How do students engage with AMR-related content on social media?
3. Which elements of the intervention were well received and how can others be improved?

## Methods

### Study Design

This study employed a mixed-methods approach to evaluate the impact of a three-month Instagram-based intervention aimed at improving undergraduate students’ knowledge, attitudes and intentions regarding AMR. The study consisted of three phases (Fig 1): (a) co-design and development of the intervention, (b) implementation of the intervention with pre- and post-intervention surveys and (c) a focus group conducted six months post-intervention to assess the impacts of the social media intervention.

**Fig 1.**
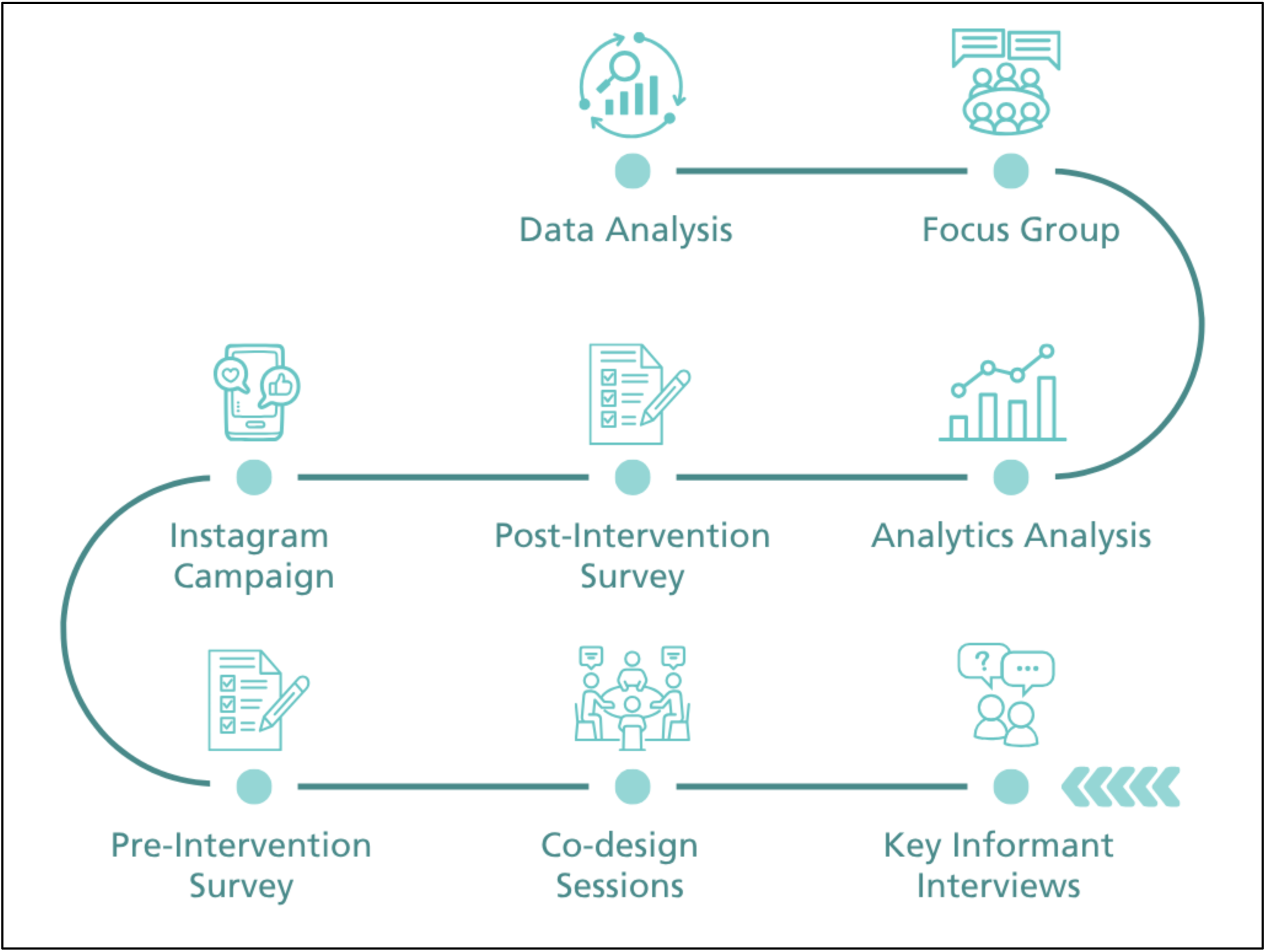
Intervention design & evaluation process

### Setting and Participants

The study was conducted at the host university and targeted first- and second-year undergraduate students from diverse disciplines. Undergraduate students were selected to enable the measurement of long-term knowledge retention within the same cohort. Recruitment strategies included email invitations sent through university mailing lists, social media advertisements and direct outreach to academic departments. Eligibility criteria included enrolment as a first- or second-year undergraduate student, willingness to follow the intervention’s Instagram page and consent to participate in surveys/ group discussions (Appendix 1).

### Intervention Development: Key informant interview and co-design sessions

A participatory co-design approach was employed to develop the intervention content. This process included multiple interactive sessions with undergraduate students and began with two key informant interviews, one with medical students and one with non-science students. Each interview involved two female participants: the first included two medical students while the second involved one student studying Languages, Linguistics & Cultures and another studying Drama. Following this, two co-design workshops were conducted with undergraduate students from a range of disciplines. The first session involved 17 participants (4 males and 13 females) and the second session included 8 participants (1 male and 7 females). The in-person sessions included brainstorming exercises, content prototyping and iterative feedback loops to generate insights into preferred social media formats, content themes and engagement strategies.

During the key informant interviews and co-design sessions with the students, it was determined that most students were not aware of AMR and belonged to either the first or second stage of behaviour change as outlined in the TTM. As the students were in the early stages of the Transtheoretical Model (precontemplation and contemplation) the intervention was designed to align with cognitive processes of change such as consciousness-raising, dramatic relief and environmental re-evaluation rather than immediate behaviour change. The study focused on influencing key cognitive precursors to action. Therefore the number of educational posts are higher than attitude and intention category posts.

### Intervention Implementation

The intervention was implemented over a three-month period (mid-January 2024 to mid-April 2024) via the project’s Instagram account. The content was routinely posted two to three times per week covering key AMR topics such as antibiotic misuse and common infections among young adults with each post aligned to target either knowledge or attitude or intentions (Appendix 3). A variety of content formats were used, including infographics, reels (short videos), carousel posts, memes and interactive stories (such as quizzes) tailored to maximise engagement (Fig 2).

**Fig 2.**
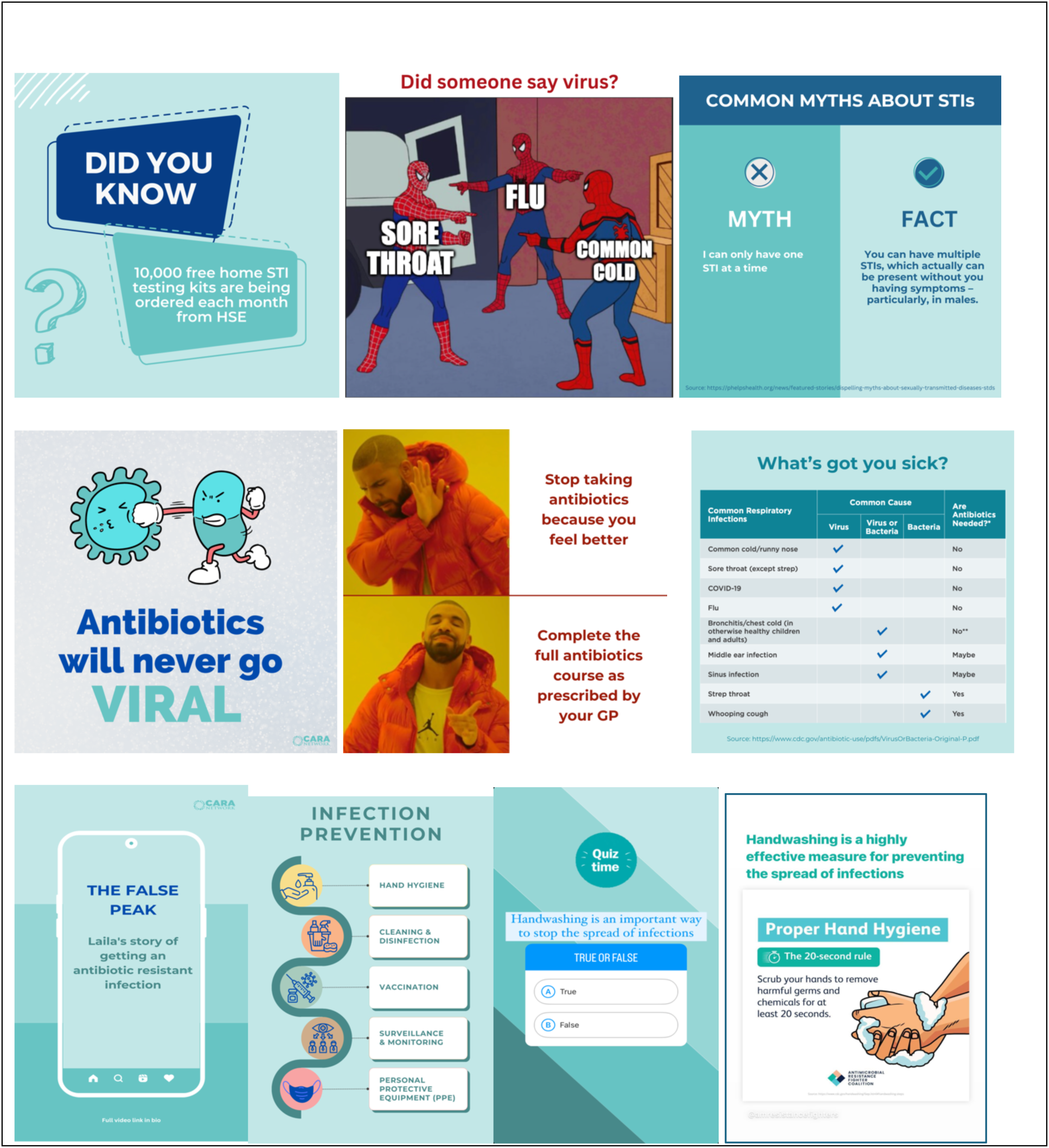
Intervention content posted on Instagram

### Evaluation

#### Pre- and Post-Intervention Surveys & Engagement metrics

Two online surveys were administered though SurveyMonkey to assess changes in students’ knowledge, attitudes and intentions regarding AMR. The questionnaire was adapted from previous AMR studies [17, 18] which aimed to measure the same parameters, to ensure reliability. The knowledge section included five validated items assessing AMR-related facts (true/false), while attitudes (3 questions) and intentions (5 questions) were measured using a 5-point Likert scale (Table 1). The survey was pre-tested with six undergraduate students to ensure the clarity and understandability of the questions. The survey was conducted pre and post intervention to assess change.

**Table 1.**
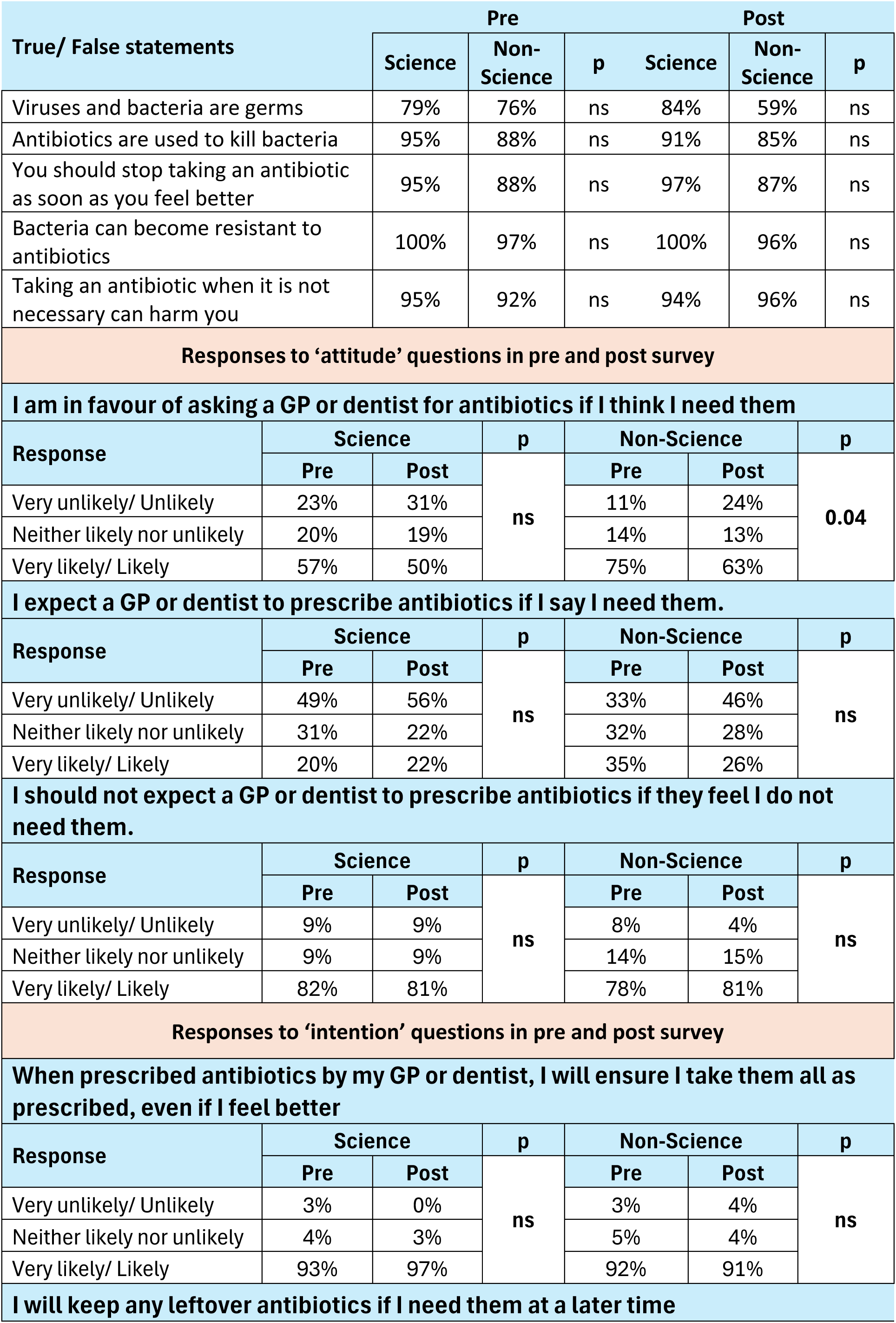
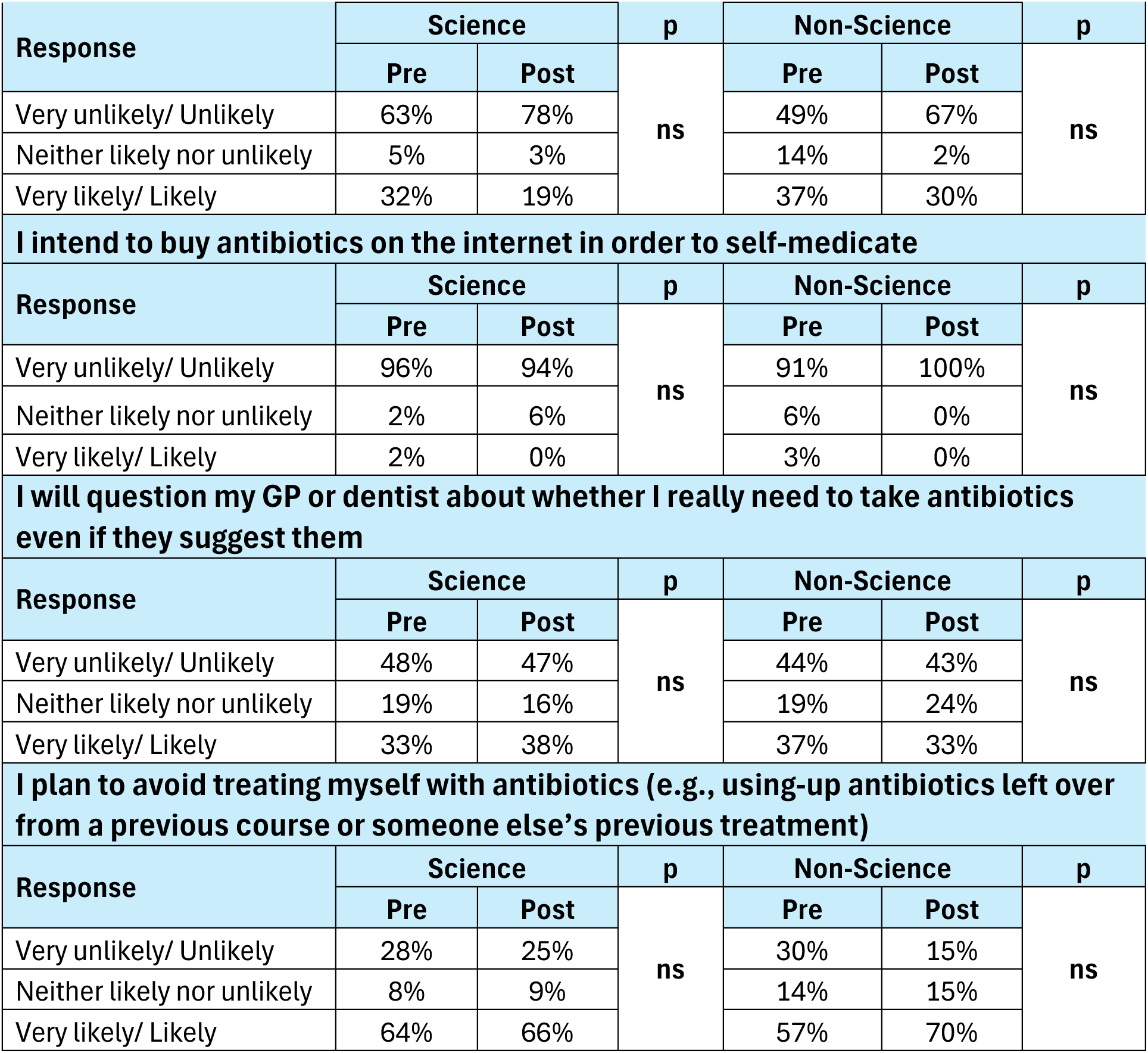
Survey Results. Percentage of correct response for Knowledge questions

The effectiveness of the intervention was also measured through Instagram engagement metrics and calculated as the sum of likes, comments, views, shares and saves divided by the follower count/reach of the post.

#### Focus group

To assess longer-term impact and knowledge retention, a focus group was conducted six months post-intervention. All students who had engaged with the intervention, answered the surveys and been a part of the co-design sessions were invited to participate. This session involved a structured discussion with students who had engaged with the intervention and included questions on engagement with content, knowledge/attitude, peer influence, social media platform effectiveness, message clarity and appeal, perceived value of the intervention, suggestions for future interventions, barriers to learning, knowledge retention and general social media use for health education (Appendix 5).

### Data Analysis

#### Quantitative Analysis

Descriptive statistics were used to summarise student distributions by gender and college. Chi-square tests were performed to assess differences in survey answers between groups. Wilcoxon signed-rank tests were performed to compare pre vs post responses on attitude and retention and the McNemar test for knowledge questions. Statistical analysis was performed in R.

#### Qualitative Analysis

Data from the co-design sessions were analysed using an inductive content analysis [19]. Two coders explored emerging themes using MaxQDA software. This dual-coder approach was used to ensure reliability, with discrepancies resolved through discussion. The analysis began with an open coding process, where data segments were systematically labelled to capture key ideas without a predefined framework. Once the initial coding was completed, codes were grouped into broader themes. The two coding versions were compared to assess consistency.

### Ethical Considerations

Ethical approval for the study was obtained from the university ethics committee. Students provided informed consent electronically before participation.

#### Patient and Public Involvement

The primary target audience of the intervention were undergraduate students who were involved from the early stages of the research. The initial key informant interviews were held with students to understand their perceptions of antimicrobial resistance (AMR), preferred social media platforms, content styles and message type that resonates with them.

Students were actively involved in the co-design of the intervention. They provided feedback on content style, tone and relevance which informed both the content development and delivery. In terms of recruitment, students advised on strategies to reach their peers including the use of university societies, student influencers and social media groups. For dissemination, students were informed that the results will be published in a scientific journal.

## Results

### Participant characteristics

A total of 400 students participated in the pre intervention survey and 78 in the post intervention-survey. Students were from a range of academic disciplines across six colleges within the university. In the pre-survey, the largest proportion of students were from the College of Business (23.2%), followed by the College of Engineering & Architecture (19%) and the College of Health & Agricultural Sciences (17.8%). The Colleges of Science and Social Sciences & Law each accounted for 14.5%, while the College of Arts & Humanities contributed 11%.

In the post-survey, the distribution changed slightly and the College of Engineering & Architecture had the highest representation (26.9%), followed by College of Science (25.6%), College of Arts & Humanities and College of Health & Agricultural Sciences (both 15.4%). Fewer students came from the College of Social Sciences & Law (11.5%) and the College of Business (5.1%).

Disciplines were grouped into ‘Science’ (College of Science and the College of Health & Agricultural Sciences) and ‘Non-Science’ (all other colleges). In the pre-survey, 32% of respondents were from Science disciplines and 68% were from Non-Science backgrounds, which changed to 41% Science students and 59% Non-Science students in the post survey.

### Key informant interview and co-design sessions

The students expressed their preference for health content on social media using formal language, having visual appeal and content clarity with interactive features. One student said *“keep them simple, bright and contrasting colours, keep text minimal”* which others agreed with. The students showed preference for content that combined short length with visual elements including infographics, reels and memes which displayed key terms in an easy-to-understand format that users could share. Students preferred content with formal language because it established trust and credibility. The combination of visual clarity and attractive design elements including bright colours, attention-grabbing hooks and minimal external links proved essential for capturing attention. The students showed strong preference for interactive content such as quizzes and stories because they wanted dynamic experiences that involved participation: *“Oh, yeah. Quizzes would be fun. I would do it”*.

Students emphasised the need to work with influencers and suggested using content formats such as carousel posts and real-life or personal stories to connect with the audience more. The students viewed educational posts as effective when they used appropriate combinations of storytelling, humour or fear-based appeals: “*So just like engage them with humour or animations by also telling them facts with human voiceovers”.* Collaborating with healthcare professionals to add credibility to sensitive topics emerged as a common point of discussion *“it’s authority. It’s medicine. It’s they know what they talk about, I would definitely watch it (GP video)”.* The sessions demonstrated that undergraduate students want educational content which combines visual appeal with credibility and diversity and interactive elements for engaging them effectively.

### Survey results

#### Overall comparison between pre-and post-survey

##### Knowledge

Science students performed better on the knowledge questions compared to non-science students before the intervention (Table 1) and this remains the same post-intervention except for the question on taking an antibiotic when it is not necessary as Non-science students have improved on their knowledge on this question. However, these differences are only significant for some questions. When comparing the knowledge of science students pre and post, there is little in the difference and none of the differences are significant. For non-science students, they actually do not improve their knowledge after the intervention, except for when to stop taking antibiotics, but none of the comparisons reach significance. Science students generally have more knowledge about antibiotics compared to non-science students, but the intervention does not significantly increase either group of students. (Table 1).

##### Attitudes

Science students are less likely to ask for an antibiotic or expect and antibiotic, compared to non-science students but they have similar attitudes not to expect an antibiotic when the GP or dentist say they do not need them. The two differences in attitude are only significant before the intervention and show that non-science students improved their attitude. (Table 1).

##### Intentions

Pre-survey, science students are less likely to keep their leftover antibiotics and planning to use these leftover antibiotics, while there are not significant differences for the other intentions. The differences in intentions between science and non-science students are not significant anymore after the intervention as non-science students improve in their intentions. Comparing before and after the interventions does not show any significant differences. (Table 1).

#### Before and after the intervention in the matched sample

A total of 47 students could be matched pre and post-intervention survey and no statistically significant differences between pre and post knowledge, attitudes or intentions was observed.

#### Instagram analytics

The Instagram account had 130 followers during the intervention. The overall engagement (the number of times students liked, commented, viewed, shared or saved a post) was 398% and knowledge posts were particularly effective, achieving a 221% engagement, followed by attitude (87%) and intentions (62%).

The intervention achieved an overall engagement of 2% by reach (how many students saw the post), with the highest engagement in the intentions category at 15%. Knowledge-related posts saw a 4% engagement, while attitude-related posts had the lowest engagement at 2% (Table 2).

**Table 2.**
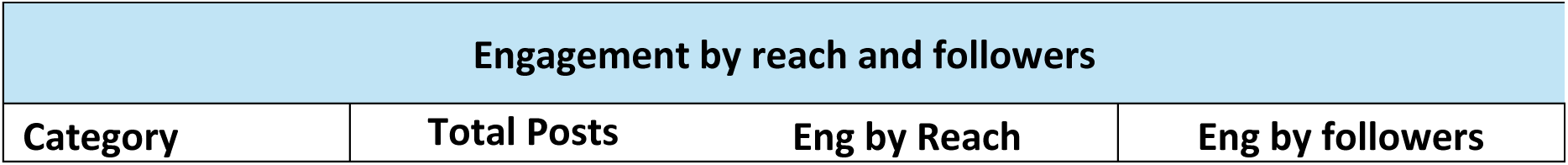
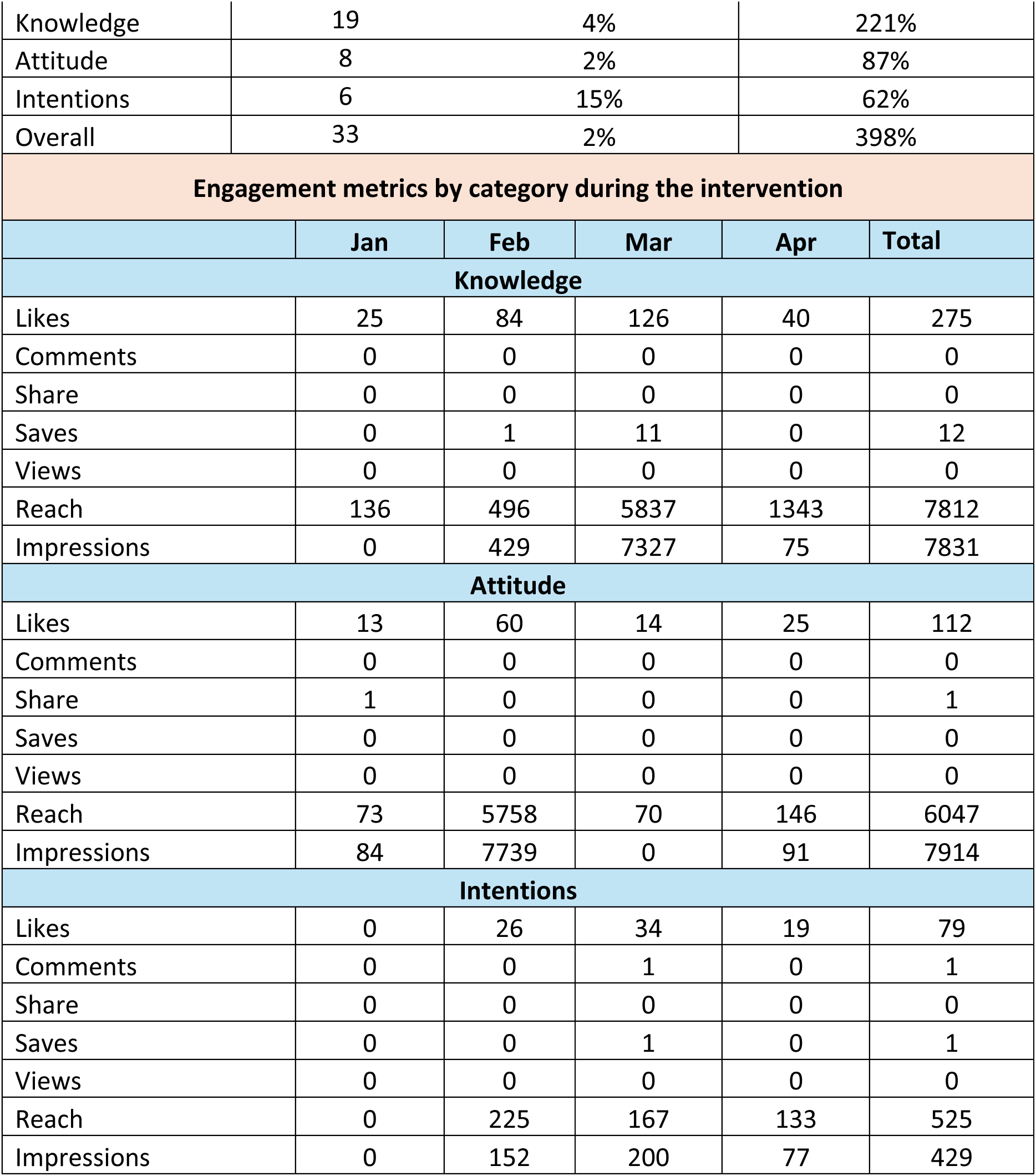
Summary of Engagement.

| Table 2 – Summary of Engagement |  |  |  |
| --- | --- | --- | --- |
| Engagement by reach and followers |  |  |  |
| Category | Total Posts | Eng by Reach | Eng by followers |

|  |  |  |  |  |  |
| --- | --- | --- | --- | --- | --- |
| Knowledge | 19 | 4% | 221% |  |  |
| Attitude | 8 | 2% | 87% |  |  |
| Intentions | 6 | 15% | 62% |  |  |
| Overall | 33 | 2% | 398% |  |  |
| Engagement metrics by category during the intervention |  |  |  |  |  |
|  | Jan | Feb | Mar | Apr | Total |
| Knowledge |  |  |  |  |  |
| Likes | 25 | 84 | 126 | 40 | 275 |
| Comments | 0 | 0 | 0 | 0 | 0 |
| Share | 0 | 0 | 0 | 0 | 0 |
| Saves | 0 | 1 | 11 | 0 | 12 |
| Views | 0 | 0 | 0 | 0 | 0 |
| Reach | 136 | 496 | 5837 | 1343 | 7812 |
| Impressions | 0 | 429 | 7327 | 75 | 7831 |
| Attitude |  |  |  |  |  |
| Likes | 13 | 60 | 14 | 25 | 112 |
| Comments | 0 | 0 | 0 | 0 | 0 |
| Share | 1 | 0 | 0 | 0 | 1 |
| Saves | 0 | 0 | 0 | 0 | 0 |
| Views | 0 | 0 | 0 | 0 | 0 |
| Reach | 73 | 5758 | 70 | 146 | 6047 |
| Impressions | 84 | 7739 | 0 | 91 | 7914 |
| Intentions |  |  |  |  |  |
| Likes | 0 | 26 | 34 | 19 | 79 |
| Comments | 0 | 0 | 1 | 0 | 1 |
| Share | 0 | 0 | 0 | 0 | 0 |
| Saves | 0 | 0 | 1 | 0 | 1 |
| Views | 0 | 0 | 0 | 0 | 0 |
| Reach | 0 | 225 | 167 | 133 | 525 |
| Impressions | 0 | 152 | 200 | 77 | 429 |

The knowledge posts received 275 likes and 12 saves but no comments, shares or views were recorded (Table 2). The total reach for knowledge was 7,812 with 7,831 impressions. March was the peak month, contributing 5,837 to the total reach and 7,327 impressions.

Attitude posts achieved 112 likes and a single share but no comments, saves and views. The reach for attitude was 6,047 and impressions totalled 7,914. The majority of this reach (5,758) and impressions (7,739) occurred in February indicating strong but short-lived engagement (Table 2).

Intentions posts received 79 likes, 1 comment and 1 save but no shares or views (Table 2). It generated a relatively lower reach of 525 and 429 impressions with February showing the highest reach (225) and March the most impressions (200).

Knowledge posts had the highest engagement and maintained consistent reach and impressions throughout the intervention period. Attitude posts had a strong engagement spike in February, suggesting a possible link to the paid campaign during this time. While intentions posts had lower overall reach and impressions, the higher engagement by reach (15%) indicates that those who saw this content were more likely to engage with it.

### Mini focus group

Eight students participated in the mini focus group session comprising one male and seven female participants. One student was studying Pharmacy while the remaining participants were from non-science disciplines. The inductive content analysis showed six major themes were identified (Figure 3).

**Figure 3:**
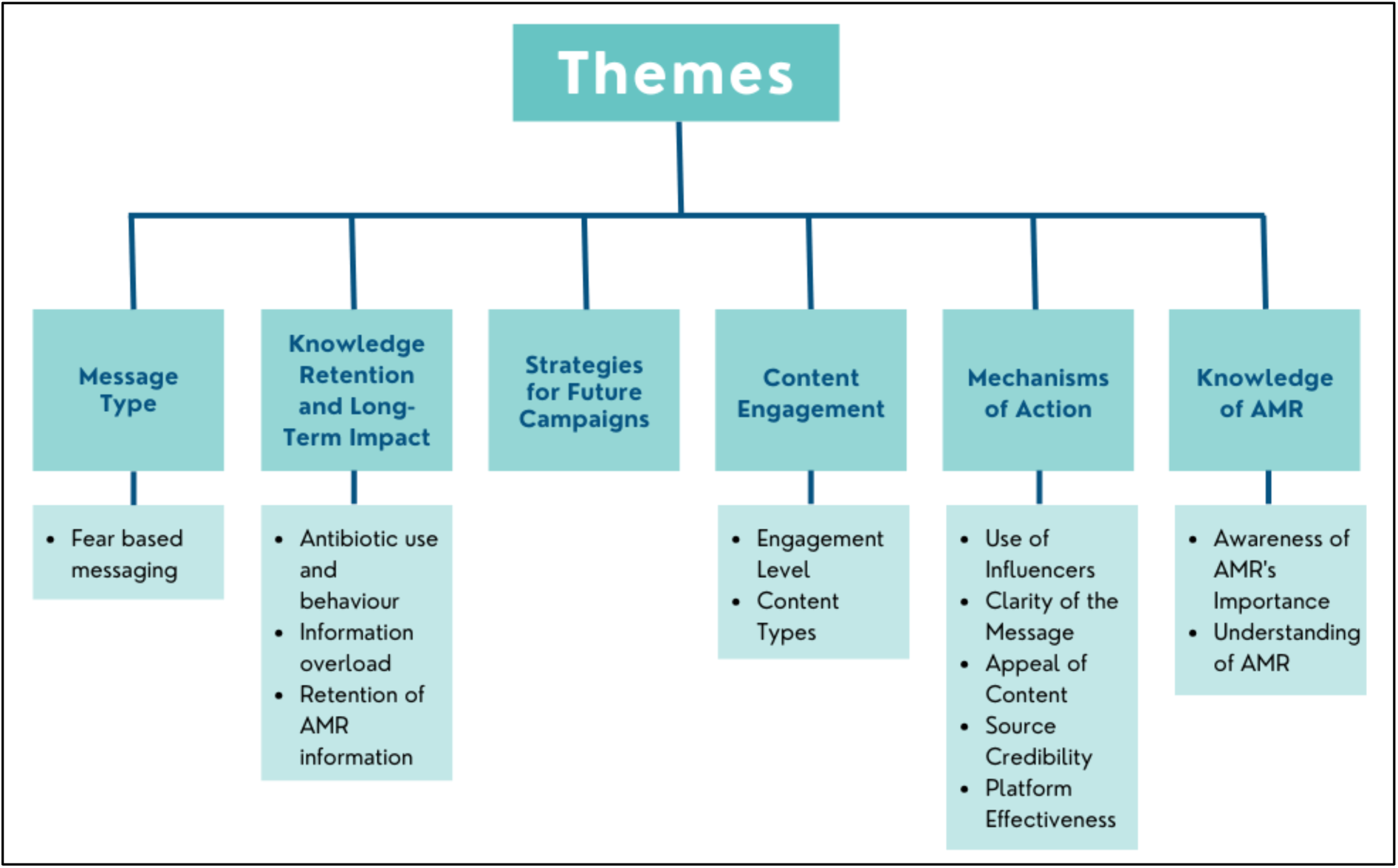
Themes from content analysis

Students indicated that fear-based messages were more memorable and effective in drawing attention to the severity of antimicrobial resistance (AMR). Statements such as, “*I think I’ll only remember it if it strikes me or makes me anxious or scared*” suggest that fear-based appeals may be instrumental in enhancing knowledge retention. Additionally, framing AMR as a “common enemy” was discussed as a potential strategy to foster collective concern and drive behavioural change in the long run.

Knowledge retention emerged as a critical concern. Students emphasised that information overload, particularly from digital platforms, posed challenges to long-term retention. It was suggested that periodic exposure such as monthly reminders would be more effective in sustaining AMR-related knowledge. Additionally, multiple students cited visual storytelling, such as documentaries or patient narratives, as more impactful than static messages.

There was consensus in influencers as key dissemination agents, yet scepticism remained about their credibility. Interestingly, students indicated that science influencers were often less effective in reaching general audiences with greater trust being placed in lifestyle and food influencers.

The effectiveness of platforms for AMR communication was another area of contrast. Some students emphasised traditional forms such as television ads, posters and flyers, whereas others highlighted digital engagement through interactive content, online ads and short-form videos. Students noted that exposure to repeated content (e.g. unskippable social media ads or flyers) played a significant role in retention. However, concerns about “mindless scrolling” on platforms such as TikTok and Instagram suggested that simply placing content online was insufficient and attention-grabbing and easily digestible formats were deemed crucial.

#### Integration with TTM Framework

The intervention was designed to engage students at the earlier stages of behaviour change particularly those in Precontemplation and Contemplation with the goal of activating cognitive and affective processes of change. These include Consciousness Raising, Dramatic Relief, Environmental Re-evaluation and Self-reevaluation which are basis to raise awareness, emotional engagement and early motivation toward change.

Thematic content analysis of the focus group data showed that the intervention activated several of these processes. For example, Consciousness Raising was evident in students’ increased understanding of AMR risks and the role of antibiotic misuse. Dramatic Relief was reflected in participants’ stronger emotional responses to fear-based and visually impactful messaging. Self-reevaluation was reflected on their own behaviours and roles in contributing to AMR.

The analysis focuses on the extent to which specific processes of change were supported rather than evaluating progression across TTM stages. This approach allows for a more precise understanding of the intervention’s cognitive impact and aligns with the theoretical positioning that different processes are relevant at different points along the change journey.

## Discussion

This study aimed to evaluate the impact and feasibility of a three-month Instagram-based health intervention designed to raise knowledge levels and influence attitudes and intentions related to antimicrobial resistance (AMR) among undergraduate students using the Transtheoretical Model (TTM) framework. The intervention specifically targeted students at earlier stages of change i.e. precontemplation and contemplation where cognitive processes are most relevant.

Qualitative findings from the focus group provided critical context and explanation, the quantitative data showed no consistent statistically significant change in knowledge, attitudes or intentions of the students (n = 47). However it is encouraging to see change in attitudes of non-science students which could be directly linked to a meme posted during the intervention. This shows students may identify meme as a way of sharing/ learning. Although, considering the numbers involved, no firm conclusions can be made.

Engagement levels with the intervention content were modest supporting the feasibility of using Instagram. Knowledge posts received higher engagement than attitudes or intentions posts, this may partially reflect content distribution as more knowledge-based posts were shared. Focus group findings indicated that students valued content that was visually engaging, used simple language, minimal text and was emotionally resonant. Memes and personal stories were particularly well received. Students also suggested improvements for future interventions including more interactive content (e.g. polls or quizzes), multiple exposure of messages and having a multi-channel strategy to ensure long-term retention.

The content analysis showed specific core cognitive and emotional processes were to a moderate degree activated including consciousness raising, dramatic relief and self-reevaluation. This finding suggests the intervention did effectively improve the level of awareness and generate certain affective responses as noted in the focus group. These findings align with the previous research that does highlight early-stage processes as important for the establishing of motivation for health behaviour change [15]. Students’ comprehension showed they understood the dangers of AMR. However, students did not fully commit themselves to enacting change. As an example, many students request antibiotics “just in case,” or rely upon passive exposure to information. These actions reflect intentions without any concrete follow-through hallmarks shown in the preparation stage.

### Comparison with prior work

Consistent with prior research the finding that fear-based or emotionally striking messages increased recall aligns with established evidence [20–22] which also shows that fear appeals can be effective. Dramatic relief has been shown to prompt intent to change, especially during early stages of change [15].

The students in our study cited information overload and short attention span as major obstacles to learning on Instagram as reported in previous studies [23, 24]. Users experience information overload when they receive more information than their processing abilities allow which results in confusion, stress and fatigue [25, 26]. This phenomenon occurs frequently on social media platforms such as Instagram as high volume of content is quickly consumed by users due to the personalised content recommendations [27]. Our study findings support these concerns about digital health content being easily scrollable but not easily remembered/ retained.

The results also confirm earlier research which shows that people trust influencers more than health institutions when it comes to health advice which highlights the growing role of peer-based credibility [28, 29]. However, students emphasised GPs being the most reliable source of health information similar to an earlier study [29].

Unlike prior studies that highlight gamified content and interactive features (like quizzes) as effective for engagement and learning [30, 31], students in our study reported minimal engagement with Instagram quizzes and little retention indicating the design of a quiz may also be an important factor.

Passive health messaging without personal relevance has been shown to be ineffective because people dismiss generic health advice [32, 33]. The effectiveness of health messages increases when they are tailored to personal relevance as it grabs attention and enhances information processing depth and learning [34]. Conversely, this study showed that passive exposure to repeated messages through physical space materials such as flyers and posters does lead to subconscious learning and may reflect the university environment. Though students spend a majority of their time at school or college, their recruitment is a challenge in research studies as highlighted in previous research [35].

Our study confirms previous research by showing that awareness by itself does not lead to changes in attitudes or intentions or small behavioural modifications such as expecting antibiotic. This discrepancy can be associated with ‘low perceived susceptibility’ as outlined in the Health Belief model [36] i.e. how likely a person believes they are to get the condition. AMR lacks the acute, personal threat often required to activate full behavioural commitment especially among young adults.

### Implications of this study

Our study highlights a co-occurrence between barriers to learning and social media platform effectiveness. Students reported that issues such as algorithmic limitation and information overload affected their ability to engage with AMR content effectively. These findings demonstrate that awareness-raising alone is insufficient. The initial development of understanding and motivation through the early-stage processes of change must be supported by mechanisms which enhance commitment, habit formation and environmental reinforcement.

### Limitations

The study used questionnaire-based quantitative data yet the main results stem from qualitative findings, which was done with a limited number of students who may not reflect the opinions of the wider population. The intervention’s exclusive use of Instagram restricts its ability to reach different demographic groups who do not actively use the platform.

### Future Research Directions

Future research needs to develop blended interventions combining digital messaging with offline reinforcement strategies which support students through different stages of change. Future interventions need to focus more on developing strategies that enhance later-stage behavioural processes to achieve lasting health behaviour change and maximise long-term impact.

## Conclusion

This study provides initial evidence that Instagram can be a feasible platform for delivering AMR-related educational interventions particularly for engaging university students. A multi-channel strategy that uses social media to grab attention along with scheduled reminders and influencer collaboration for peer-driven engagement would improve intervention effectiveness.

## No competing interests

All authors have completed the ICMJE uniform disclosure form and declare: no support from any organisation for the submitted work; no financial relationships with any organisations that might have an interest in the submitted work in the previous three years; no other relationships or activities that could appear to have influenced the submitted work.

## Appendices

Appendix 1 - Consent forms, PIL, recruitment emails, recruitment poster

Appendix 2 - Key informant interview questions and co-design questions

Appendix 3 - All Instagram posts and engagement

Appendix 4 - Questionnaire - pre and post

Appendix 5 - Focus group questions

## Data Availability

All data produced in the present study are available upon reasonable request to the authors

